# The impact of respiratory infections and probiotic use on the nasal microbiota of frail residents in long-term care homes

**DOI:** 10.1101/2023.06.02.23289167

**Authors:** DME Bowdish, L Rossi, MB Loeb, J Johnstone, LP Schenck, M Fontes, MG Surette, FJ Whelan

## Abstract

**Background:** Residents in long-term care (LTC) homes, who tend to be of advanced age and frail, are at increased risk of respiratory infections. The respiratory microbiota is known to change with age, but whether these changes contribute to the risk of infection is not known. Our goal was to determine how the nasal microbiota of frail older adults changes during symptoms of influenza-like illness (ILI) and how this may be impacted by enrollment in a placebo-controlled trial testing the feasibility of administering a *Lactobacillus rhamnosus* GG probiotic to prevent respiratory infection (2014 - 2017). The microbiome of the nasal (mid-turbinate) of 150 residents of LTC homes was interrogated using 16S rRNA gene sequencing.

**Results:** We identified a diverse and individualized microbiota which could be separated into 9 distinct clusters based on Bray Curtis distances. Samples collected during symptoms of influenza-like illness (ILI) differed statistically from those collected pre- and post-cold and influenza season, and we observed decreased temporal stability – as measured by movement between clusters – in individuals who experienced ILI compared to those who did not.

**Conclusions:** The use of probiotics decreased ILI-induced changes to the microbiota; however, it is not clear whether this decrease is sufficient to prevent respiratory illness.

## Background

The burden of respiratory infections in long-term care (LTC) residents is high^1^, and the devastating mortality and frequent outbreaks that occurred during the SARS-CoV-2 pandemic were a painful reminder that systemic features of care homes such as staffing patterns, ventilation, and crowding can be major factors in infection rates^2^, ^3^. Independent of the increase in risk associated with LTC homes, residents are still vulnerable to infection due to their advanced age, frailty, and chronic health conditions. In fact, frailty is a better predictor of infection risk and poor outcomes than chronologic age^4, 5^ likely due to the systemic inflammation and immune remodelling that occurs in frail individuals^6^.

Carriage rates of common pathogens like *Streptococcus pneumoniae* are counterintuitively reported to decrease with age, despite the fact that susceptibility to pneumococcal infection increases with age^7, 8^. It is believed that this is because pneumococcal carriage stimulates anti-bacterial immunity in the lungs; alveolar macrophages from individuals who are experimentally colonized have enhanced killing of both pneumococcus and other respiratory pathogens^9^. Age-related changes in other members of the airway microbiota have also been reported, and these may contribute to susceptibility to both bacterial and viral infections. As an example, individuals who are colonized with *Corynebacterium* spp. are less likely to naturally carry or be experimentally colonized with pneumococcus^10, 11^. Lower relative abundance of both *Moraxella* spp., and *Dolosigranulum pigrum* have been reported in children hospitalized for serious respiratory infections, but whether these are truly associated with protection from infection or whether they decrease in abundance during the course of infection is unclear^12, 13^. Similarly, older patients with pneumonia have outgrowth of some microbes in the upper respiratory tract microbiota^14^ but the degree to which this contributes to infection is not known. Age, frailty, LTC home, specific health conditions, and immune senescence have been previously shown to be associated with age-associated changes in the gut microbiota^15, 16^ but whether these factors influence the upper respiratory tract is not known. Understanding if members of the upper respiratory tract microbiota can protect against infection may provide novel preventative strategies in older, frail individuals who are the most likely to have poor outcomes resulting from respiratory infection.

In order to understand the role of the microbiota in respiratory infection in frail older adults, we analyzed samples from 150 residents of LTC homes who had been enrolled in a randomized, double-blinded, placebo-controlled clinical trial testing the feasibility of probiotics to prevent respiratory infection^17^. Samples were collected from individuals not experiencing respiratory illness at the onset of cold and influenza season (Nov-Dec), whenever a resident experienced an influenza-like illness (ILI) event, and after cold and influenza season in the absence of illness (May-June). We investigated whether frailty, health conditions, and systemic inflammation altered the composition of the nasal microbiota and whether there were features of the microbiota that predicted susceptibility to ILI. We found a diverse microbiota that could be divided into 9 clusters. The microbiota of samples collected during ILI was statistically distinct from those collected outside of illness, and – when examined longitudinally – individuals who experienced symptoms of respiratory infection experienced decreased temporal stability of their nasal microbiota then those who did not. These affects appear to be mitigated with the use of probiotics, however a larger follow up study is warranted to reach a definitive conclusion.

## Methods

### Participant recruitment and sample collection

Samples were collected from individuals (n=150) as part of a multi-site, randomized, placebo-controlled trial on the feasibility of administering probiotics to prevent respiratory tract infections in long-term care (LTC) residents^17^. Residents from 12 LTC homes in Ontario, Canada who were ≥ 65 years old were recruited over a 4-year period (2014-2017). Here, we sampled the nasal microbiota of samples collected from the later 3 years of the pilot study (**Sup Fig 1**). Participants provided a flocked nasal (mid-turbinate) swab in universal transport medium (Copan Italia, Brescia Italy) prior to cold and influenza season (Nov-Dec), when they had symptoms consistent of an influenza-like illness (ILI; as determined by a trained nurse; cough, nasal congestion, sore throat, headache, sinus problems, muscle aches, fatigue, earache or infection, chills) and following the end of cold and influenza season (May-June). Exclusion criteria included residents on immunosuppressive drugs, who had hematological malignancy, structural heart disease, gastroesophageal or intestinal injury, or individuals who were at high risk of an endovascular infection. Participants were randomized to receive a probiotic (2 capsules of *Lactobacillus rhamnosus* GG (Culturelle, CH Hansen, Hoersholm, Denmark; estimated 10 billion colony forming units (CFUs) per capsule) daily or a placebo (calcium carbonate) for 6 months. Details of probiotic administration have been previously published^17^. There were no differences in participant demographics between the placebo and probiotic groups (**Sup Table 1**; reference ^17^).

**Figure 1:**
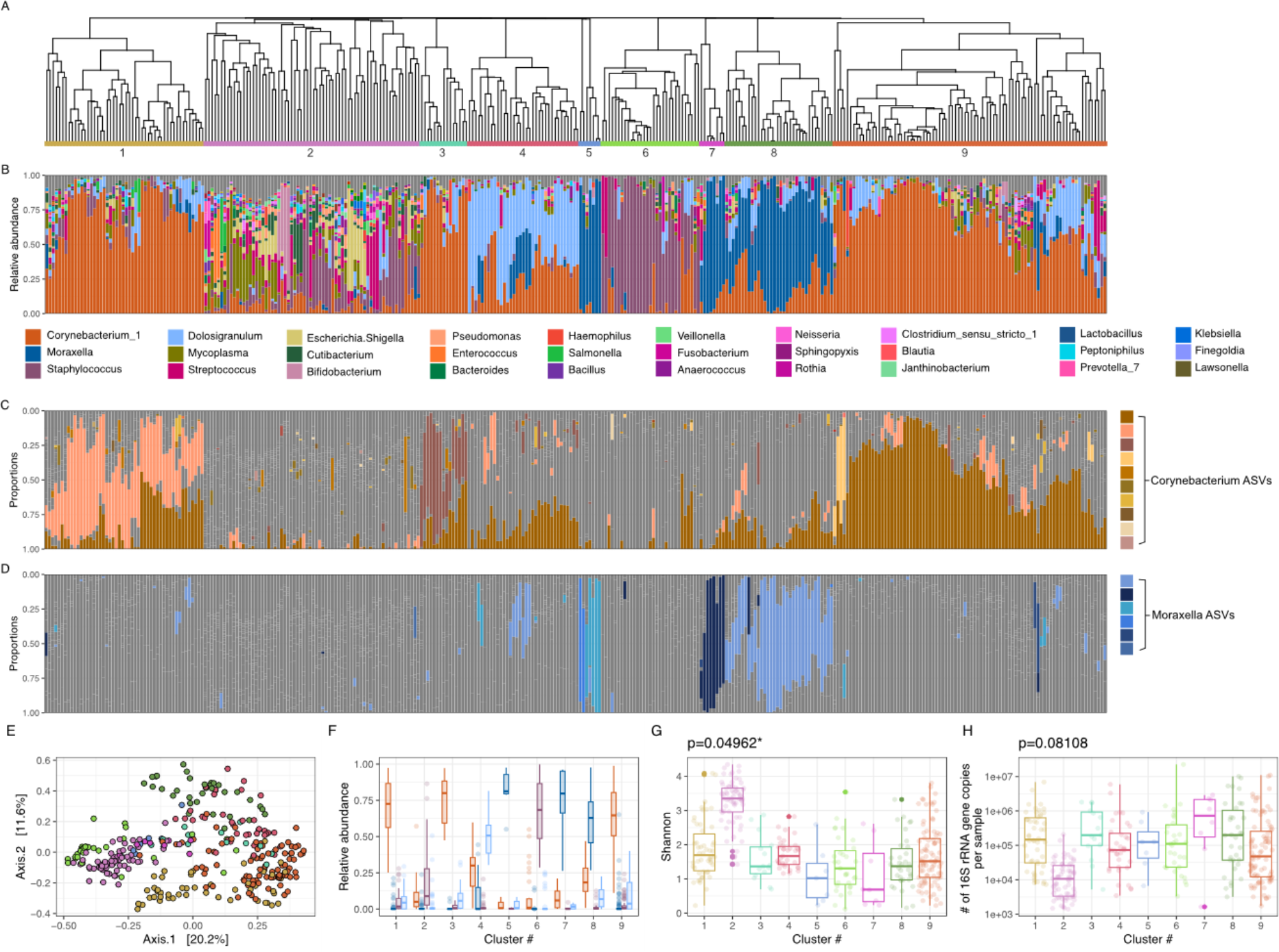
The nasal microbiota of frail older adults groups into 9 distinct clusters. A. A dendrogram based on hierarchical clustering of Bray Curtis distances between samples. Coloured and numbered bars indicate the 9 clusters determined by hierarchical clustering. **B.** Taxonomic summaries ordered according to the sample order in the dendrogram accompanied by a legend of the most abundant 30 genera. **C-D**. The relative abundance of each amplicon sequence variant (ASV) within *Corynebacterium* (**C**) and *Moraxella* (**D**). Each ASV is coloured a different shade of orange/blue; grey bars indicate relative abundances of other taxa **E.** A PCoA plot of the Bray Curtis distances of all samples within the dataset coloured by cluster membership. Colours of each cluster match those used in **panel A**. **F.** The mean relative abundances of the 4 most abundant taxa separated by cluster. Colours of each taxa match those used in **panel B**. (*Corynebacterium*: orange, *Moraxella*: dark blue, *Staphylococcus*: purple, *Dolosigraniulum*: light blue). **G.** The median Shannon diversity metric differs significantly between clusters (p=0.04962; Levene’s test); **H.** however, the median qPCR concentrations do not (p=0.08108, Levene’s test).

**Table 1:** Participant (n=150) characteristics.

| Characteristic | Properties |
| --- | --- |
| Sex | 103 Female (68.7%) |
| Median age (± standard deviation; sd) | 86.5 ($\pm 7.1$) |
| Smoker | 5 Yes, 71 No, 73 Prior (1 Unknown) |
| Median # of medications (± sd) | 9 ($\pm 3.6$) |
| Median # of co-morbidities (± sd) | 9 ($\pm 3.2$) |
| Median Barthel score (± sd) | 7 ($\pm 5.78$) |
| Num. of Individuals reporting Respiratory Events | 51 (34.0%) |
| Mean # of events per individual | 1.12 |
| Probiotic Group | 77 Active (51.3%), 73 Placebo (48.7%) |

Informed consent was received by the participants or their substitute decision makers. All protocols were approved by the Hamilton Integrated Research Ethics Board.

Of the collected samples, 334 from 150 individuals (n=150 pre-cold and influenza season, 57 ILI, 127 post-cold and influenza season) passed stringent quality control measures including verification of the 16S rRNA gene PCR product on an agarose gel, quantification of 16S rRNA gene DNA load, and a minimum number of high-quality DNA sequencing reads (see below for more detail).

### DNA extraction and amplification of the 16S rRNA gene

DNA extraction was performed as previously described^18^. 300 μl of sample was resuspended in 800 μl of 200 mM NaPO4, 100 μl of guanidine thiocyanate- ethylenediaminetetraacetic acid-Sarkoyl, and together homogenized using 0.2g of 0.1- mm glass beads (Mo Bio, Carlsbad, CA). 50 μl of lysozyme (100 mg/ml), and 10 μl RNase A (10 mg/ml) were added to the sample and incubated at 37°C for 1 hour to enzymatically lyse the sample. Following, 25 μl of 25% sodium dodecyl sulfate, 25 μl proteinase K, and 62.5 μl 5 M NaCl were added and incubated at 65°C for 1 hour. Samples were then subject to centrifugation at 12,000 x g. The supernatant was subsequently removed to a new microcentrifuge tube to which an equal volume of phenol-chloroform-isoamyl alcohol was added and the sample again centrifuged. The solution with the lowest density was transferred to a new microcentrifuge tube and 200 μl of DNA binding buffer (Zymo, Irvine, CA) added. The solution was then transferred to a DNA column (Zymo), washed, and DNA eluted using ultrapure H2O.

Following, amplification of the 16S rRNA gene variable 3 (v3) region was performed as previously described^19^ with some modifications. 341F and 518R 16S rRNA gene primers were adapted to the Illumina (San Diego, CA) platform with the inclusion of unique 6-base pair barcodes to the reverse primer to allow for multiplex amplification^19^. A 50 μl PCR reaction was performed in three equal volume reactions, collectively containing 5 pmol of each primer, 200 μM of each deoxynucleoside triphosphate (dNTP), 0.4 mg/mL BSA, 1.5 mM MgCl2, and 1 U *Taq* polymerase (Life Technologies, Carlsbad, CA). The PCR reaction was subject to an initial denaturation step at 95°C for 5 min followed by 35 cycles of 95°C for 30 sec, 50°C for 30 sec, and 72°C for 30 sec; the incubation ended with an extension step at 72°C for 7 min. The presence of a PCR product was verified by electrophoresis (2% agarose gel) and only those samples with visible bands were sent for normalization using the SequelPrep kit (ThermoFisher, #A1051001) and DNA sequencing on the Illumina MiSeq platform. A positive control sample of known community composition sequenced in parallel to these data contained the same 50 ASVs in similar proportions as the positive control samples run on prior and subsequent MiSeq runs. Four negative controls – including DNA extraction and PCR controls – resulted in <1520 bacterial reads per sample, none of which were consistently assigned to the same ASVs. All raw sequencing data is available on NCBI’s SRA PRJNA858212.

### Processing of 16S rRNA gene sequencing data

Raw reads were initially processed with Cutadapt^20^ to trim the adapter and PCR primer sequences and filter to a minimum quality score of 30 and a minimum length of 100bp. DADA2^21^ was used to resolve sequence variants for results from each separate Illumina run prior to merging data from all runs together. Amplicon sequence variants (ASVs) were then filtered for bimeras; taxons were classified using the SILVA database version 1.2.8^22^.

### Quantification of 16S rRNA gene DNA load via qPCR

Because samples from the nose have low microbial concentrations, we assessed the extracted DNA via qPCR in order to quantify the number of copies of 16S rRNA gene present in each sample. The protocol was adapted from^23^; briefly, reactions were carried out in a 96-well plate in a 20 μL mixture containing 10 pmol of forward (926F AAA CTC AAA KGA ATT GAC GG) and reverse (1062R CTC ACR RRC ACG AGC TGA C) primer^24^, 1 μL of extracted swab DNA, 10 μg of bovine serum albumin, water, and Eva SsoFast EvaGreen supermix (Bio-Rad, Canada). Samples were placed in a Bio-Rad CFX96 Thermocycler (Bio-Read, Canada) and were subject to an initial denaturing step (98C for 2 minutes), followed by 40 cycles of 5 seconds at 98C and 5 seconds at 60C. Melt curve analysis was generated by 0.5C increments for 5 seconds from 65C to 95C to ensure the generation of a single PCR product. Each reaction was performed in triplicate, with cycle thresholds converted to copies of 16S rRNA gene via standard curve of known quantities of *Escherichia coli* DNA within each qPCR plate.12 samples which had <10^3^ copies of 16S rRNA gene sequence per sample were removed from all subsequent analyses.

### Statistical analyses of 16S rRNA gene sequencing data

The above quality control measures resulted in a total of 334 samples included in microbiome analyses. 39 metadata data points were collected; to avoid over-interpreting any correlations of such data with microbial composition, metadata variables were only considered if: (a) σ:15% of the data points were unknown/missing; (b) for binary variables, there was ≥10% variation; (c) for non-binary discrete variables, each value accounted for >3% of the overall variation (otherwise, the value was omitted). All continuous variables were included.

To identify any possible correlations between metadata variables and avoid reporting any indirect associations between metadata and microbial composition, each pair of variables were investigated using a chi-squared or aov test (depending on the data type). When the statistical test resulted in a p-value <0.05, we rejected the null hypothesis that the variables tested were independent. A list of all correlating variables is included in **Sup Table 2**.

**Table 2:** p- and R^2^ values of anova statistical test of association between metadata variables and the composition of the microbiome.

| Characteristic | By sample distance metrics (Bray Curtis p-value ($R^2$ value)/Aitchison p-value ($R^2$ value)) | By cluster membership (test statistic in brackets) |
| --- | --- | --- |
| Age at enrolment | 0.224(0.008)/0.821(0.006) | 0.295 (aov) |
| Sex | **0.047\*(0.013)**/0.149(0.008) | 0.253 (aov) |
| LTC home site | **0.033\*(0.082)**/0.084(0.068) | 0.616 (chisq) |
| Month | 0.699(0.018)/**0.001\*\*\*(0.034)** | 0.745 (chisq) |
| Season | 0.966(0.003)/**0.004\*\*(0.011)** | 0.747 (aov) |
| Year | 0.204(0.017)/**0.001\*\*\*(0.032)** | 0.650 (aov) |
| Clinical trial treatment group (i.e., probiotics or placebo) | 0.921(0.004)/0.439(0.007) | 0.832 (aov) |
| Had respiratory event | 0.402(0.007)/0.079(0.008) | 0.120 (aov) |
| Smoker | 0.782(0.017)/0.367(0.021) | 0.390 (aov) |
| Medications (number of) | 0.346(0.007)/0.859(0.006) | 0.674 (aov) |
| Influenza vaccination (current season) | 0.837(0.004)/0.307(0.007) | 0.703 (aov) |
| Influenza vaccination (previous season) | 0.261(0.015)/0.358(0.014) | 0.399 (aov) |
| Influenza vaccination (ever) | 0.095(0.019)/0.175(0.015) | 0.339 (aov) |
| Pneumococcal vaccination (ever) | 0.689(0.006)/0.586(0.007) | 0.901 (aov) |
| Shared room (yes/no) | 0.480(0.006)/0.370(0.007) | 0.490 (aov) |
| Barthel total | 0.121(0.01)/**0.003\*\*(0.011)** | 0.653 (aov) |
| COPD | 0.059(0.011)/0.441(0.007) | 0.911 (aov) |
| CHF | 0.365(0.007)/0.825(0.006) | 0.651 (aov) |
| CVD | **0.017\*(0.015)**/0.083(0.009) | 0.477 (aov) |
| Anemia | 0.923(0.003)/0.673(0.006) | 0.977 (aov) |
| Dementia | 0.912(0.004)/0.597(0.006) | 0.449 (aov) |
| Stroke | 0.074(0.011)/0.449(0.007) | 0.203 (aov) |
| Diabetes mellitus | 0.108(0.01)/0.191(0.007) | 0.104 (aov) |
| Hypothyroid | 0.957(0.003)/0.863(0.006) | 0.998 (aov) |
| Comorbidities (number of) | 0.765(0.005)/0.337(0.007) | 0.891 (aov) |
| Seizures | 0.555(0.006)/0.609(0.006) | 0.822 (aov) |
| Cancer | 0.531(0.006)/0.422(0.007) | 0.385 (aov) |
| IL-1β | 0.887(0.004)/0.830(0.005) | 0.483 (aov) |
| IL6 | 0.866(0.004)/0.921(0.005) | 0.831 (aov) |
| TNFA | 0.652(0.006)/0.990(0.004) | 0.177 (aov) |

**COPD: chronic obstructive pulmonary disease, CHF: congestive heart failure, CVD: cardiovascular disease.**

All statistical analyses were performed in R v3.6.1^25^ primarily using the phyloseq v1.28.0^26^ and vegan v2.5.6^27^ packages. Alpha diversity was calculated using the Shannon index. Beta diversity was determined using both Bray Curtis and Aitchison distances (using R’s microbiome package v1.14.0^28^ in addition to phyloseq); to calculate Bray Curtis distances, the dataset was rarefied to the minimum number of reads per sample in the dataset (n=1268). The composition of microbiome communities in relation to included metadata variables was interrogated using a permanova statistical test (adonis function of the vegan package v2.5.6^27^). Differentially abundant ASVs were determined using ANCOMBC^29^. Networks of cluster movement between pre-C&F, ILI, and post-C&F clusters were determined using R’s igraph v1.2.6 and visualized using Gephi^30^. R packages ggplot2^31^ and patchwork (https://github.com/thomasp85/patchwork) were used to generate visuals. All code is provided as a supplemental R markdown file (**Sup File 1**).

### Clustering of samples via hierarchical clustering

The Silhouette method, encoded in R’s factoextra v1.0.7 package^32^, was used to determine the optimal number of clusters for complete hierarchical clustering based on the Bray Curtis distance between samples and associated PCoA scores. Using this method, the optimal number of clusters was determined to be 10; of these, one cluster had a size of n=1 and was thus excluded from future analyses. Hierarchical clustering was performed using R’s cluster v2.1.0 package^33^. The resulting 9 clusters containing 334 samples were tested with the vegan package for statistically significant dispersion (betadisper^27^; p=<2.2e-16), differences between cluster centroids (adonis^27^; p=0.001), and to be significantly different in an analysis of similarity (anoism^27^; R=0.726, p=0.001), indicating that subsetting the data in this way generated statistically significant clusters. A dendrogram of all samples was split into 9 clusters with dendextend v1.15.1^25^ and visualized with ggtree v3.0.3^34^.

## Results

### Participant demographics

Mid-turbinate samples of the nose were collected from 150 individuals residing in LTC homes in Ontario, Canada (see *Methods*). Participants were predominantly female, with a median age of 86.5 years (**Table 1**). Individuals were on a median of 9 medications and had a median of 9 co-morbidities. Participant’s Barthel scores – an index between 0 and 20 used to describe performance in daily living^35^ – were diverse, ranging from 0 to 20 with a median value of 7 (**Table 1**). 34.0% of study participants reported symptoms of influenza-like illness (ILI), with 44 participants experiencing 1 event and 7 participants experiencing 2 events. Participants in this trial were split into daily probiotic and placebo arms of this study (**Table 1**) which were evenly matched for all participant characteristics (**Sup Table 1**).

### The composition of the nasal microbiota of long-term care residents

The nasal microbiota of LTC residing older adults is highly variable (**Fig 1A-B**). Within the 334 mid-turbinate samples analyzed, we observed 720 genera, of which 662 had a cumulative relative abundance of ≥0.01%, and 122 of which were observed at ≥1% relative abundance in ≥1 sample. On average, an individual sample contained 56 genera; however, this ranged from 7 to 146 (sd: ±27.4). *Corynebacterium* is the most abundant and most prevalent genera, with a mean relative abundance of 37.9% (median: 32.0%, range 0.04-98.7%) and being present in 332 of the 334 samples. Other abundant genera include *Moraxella* (mean relative abundance 1.18%, range 0.002-99.8%), *Staphylococcus* (mean relative abundance 1.17%, range 0.01-99.77%), and *Dolosigranulum* (mean relative abundance 1.11%, range 0.004-81.49%). The variability between individuals was large, as evidenced by a mean Bray Curtis distance of 0.80 (median: 0.87).

Because of the observed inter-individual variability, we applied hierarchical clustering on the composition of the microbiome which identified 9 clusters in our dataset (**Fig 1A**) which were verified using multiple statistical tests (see *Methods*; **Sup Fig 2A-C**). Samples from each cluster separate in a PCoA analysis across multiple axes (**Fig 1E**; **Sup Fig 2D-G**), and have more similar Bray Curtis distances within clusters than comparisons across clusters (**Sup Fig 2C,H**). The median number of samples per cluster is 34, ranging from n=7 (cluster 5) to n=86 (cluster 9). All but one cluster is associated with a dominant taxon (>35% relative abundance in >70% of samples; **Fig 1B,F**). For example, clusters 6 and 4 are associated with a high mean relative abundance of *Staphylococcus* and *Dolosigranulum*, respectively. There are 3 clusters (clusters 1,3,9) in which *Corynebacterium* is the dominant genera, but each is associated with a unique ASV (**Fig 1C**). Similarly, *Moraxella* is the dominant genera of clusters 5,7, and 8 and each cluster is associated with a particular profile of ASVs (**Fig 1D)**.

**Figure 2:**
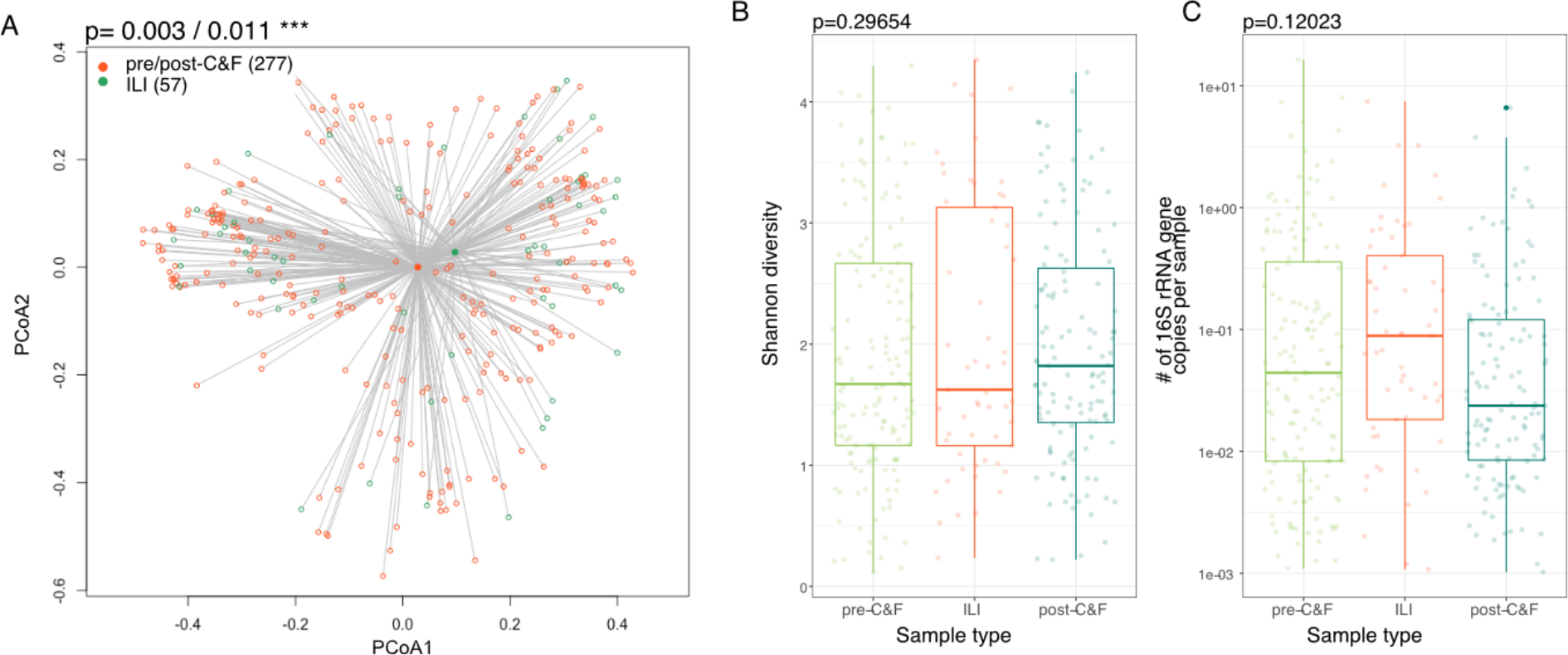
The composition, but not the α-diversity or bacterial load, of the frail older adult nasal microbiota is significantly altered during ILI events. A. Community-wide, the microbiome composition changes significantly between samples collected during ILI versus times of relative health (p=0.003/0.0011, permanova with Bray Curtis and Aitchison distances). **B-C.** However, there is no significant change in α-diversity (as measured by the Shannon metric; **B**) or bacterial load (as measured by qPCR Concentration; **C**) when samples collected during ILI were compared to those collected pre- and post-cold and flu season. pre-C&F = pre-cold and flu season; ILI = influenza like illness; post-C&F = post-cold and flu season.

Uniquely, cluster 2 is not associated with a particular dominant taxon (**Fig 1F**) and is the most diverse (Shannon index, **Fig 1G**), and shared the least Bray Curtis similarity across samples (**Sup Fig 2H**). We hypothesized that the absence of a dominant taxon might mean a decrease in the total bacterial load. When we quantified total microbial DNA by qPCR, we did not find a statistically significant difference in bacterial DNA between clusters although the median value was lower in cluster 2 than in any other cluster (**Fig 1H**). However, when we investigated whether bacterial load significantly correlated with any particular ASV(s), we did not find a correlation with any dominant taxa. Instead, decreasing microbial load was associated with increased levels of other organisms also commonly associated with the microbiome of the oral cavity and skin (e.g., *Streptococcus* and *Cutibacterium,* respectively; **Sup Fig 3**). We hypothesize that this may mean that in samples with a low bacterial load, the unique biogeography of the mid-turbinates is lost as we have previously shown in the nasopharyngeal microbiome of frail older adults^18^.

**Figure 3:**
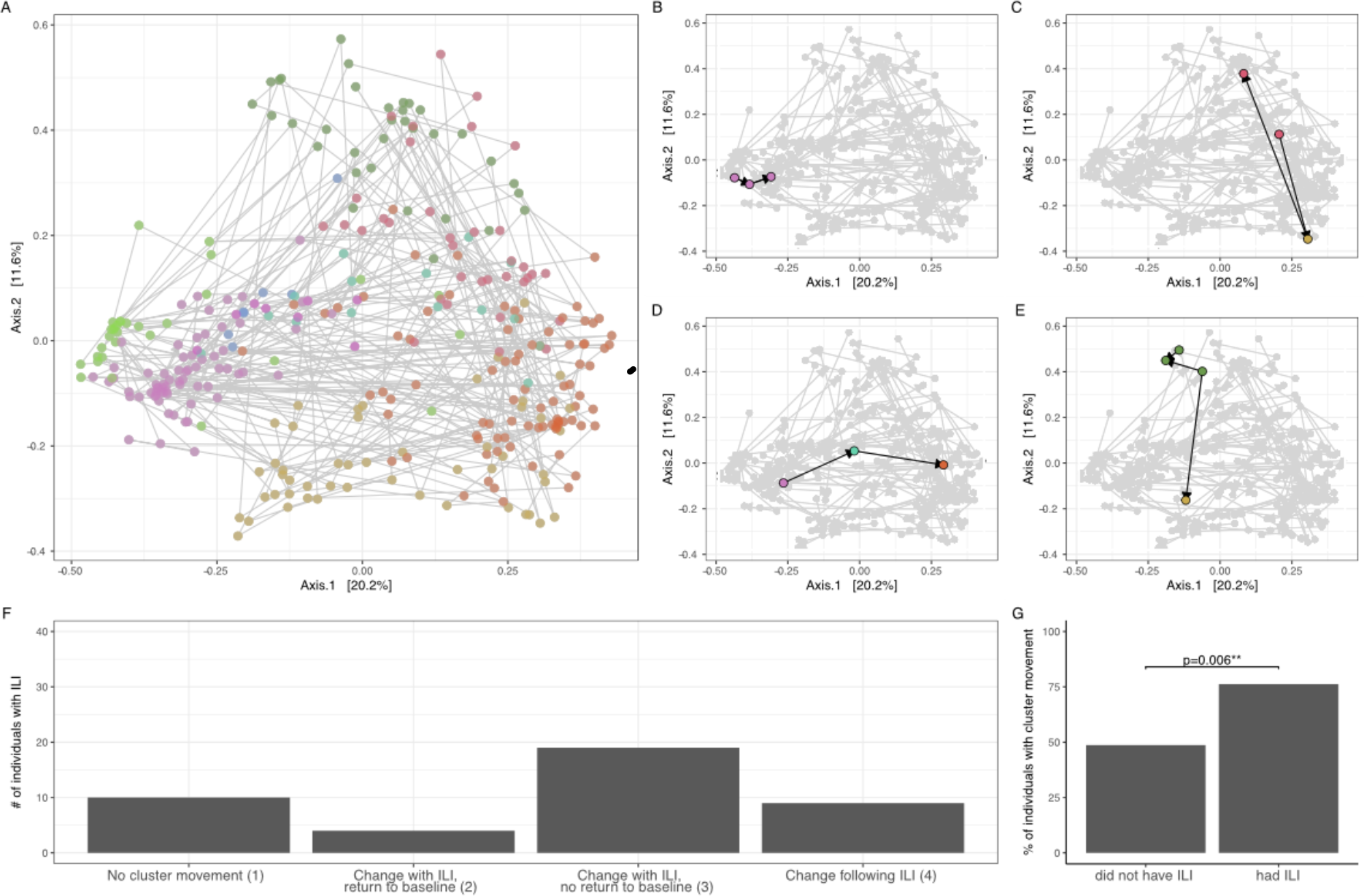
The longitudinal effect of ILI on the frail older adult nasal microbiota. A. The PCoA of all samples coloured by cluster with lines connecting longitudinal samples of each individual. Examples of individuals which stay in the same cluster throughout sampling (**B**, category 1), change cluster upon ILI but return to their pre-ILI cluster upon resolution (**C**, category 2), change cluster upon ILI but do not return to their original cluster (**D**, category 3), and change cluster following an ILI (**E**, category 4). Counts of individuals who fall into each of these cluster movement categories are quantified (**F**) and the frequency of cluster movement is compared to that of individuals who did not experience respiratory events (**G;** p=0.006, chi-squared test).

### Participant characteristics and relationship to the composition of the airway microbiota

We tested 30 metadata variables (**Table 2**), 5 of which (sex, LTC home site, time of collection, frailty, cardiovascular disease) were significantly correlated with the composition of the mid-turbinate microbiota against either of two β-diversity metrics employed (p:<0.05, PERMANOVA using either Bray Curtis or Aitchison distance; **Table 2** see *Methods*). However, none of these 5 variables were significant across both β- diversity distance metrics and could each only explain <5% of the observed variance in the data (**Table 2**); in contrast 68.84% of the variability in the dataset was explained by inter-individual differences.

Biologic sex significantly correlated with the composition of the microbiome (p=0.047, R^2^=0.013, PERMANOVA using Bray Curtis); however, there was no association of β- diversity or cluster membership with age. In contrast, frailty (as measured by the Barthel score) correlated with microbiota composition (p=0.003, R^2^=0.011, PERMANOVA using Aitchison). Because chronic inflammation (‘inflamm-aging’) is associated with both frailty and immune dysfunction, we investigated whether there were any relationships with circulating inflammatory mediators, specifically TNF, IL- 1β, and IL6 and chronic health conditions such as cardiovascular disease, dementia, and chronic obstructive pulmonary disease (COPD). Although there were no associations with inflammatory cytokines, community composition in individuals with cardiovascular disease was significantly different (p=0.017, R^2^=0.015, PERMANOVA using Bray Curtis).

### COPD: chronic obstructive pulmonary disease, CHF: congestive heart failure, CVD: cardiovascular disease

The composition of the microbiome differed between the 10 LTC facilities (p=0.033, R2=0.082, PERMANOVA using Bray Curtis), consistent with previous studies of the gut microbiota^16^. 9 ASVs with a mean relative abundance >0.1% were differentially abundant across LTC sites (ANCOMBC); many of these ASVs correspond to the dominant taxa in the dataset – including *Moraxella, Corynebacterium,* and *Dolosigranulum* (**Sup Fig 4A-C**). There were 8 metadata variables that correlated with LTC home site including those associated with the time of collection (i.e., month, season, and year of collection) and other variables that likely reflect differences in LTC home practices (e.g., influenza and pneumococcal vaccination, whether the resident was in a shared or private room; **Sup Table 2**). Of these, the time of sample collection also significantly correlated with microbiome composition (**Table 2**). 5 ASVs with >0.1% relative abundance were differentially abundant by month of sample collection, including the same *Moraxella* ASV which was differentially abundant by LTC home site (**Sup Fig 4D**). Collectively these data demonstrate that seasonality and LTC home site may have some effect on the mid-turbinate microbiota.

**Figure 4:**
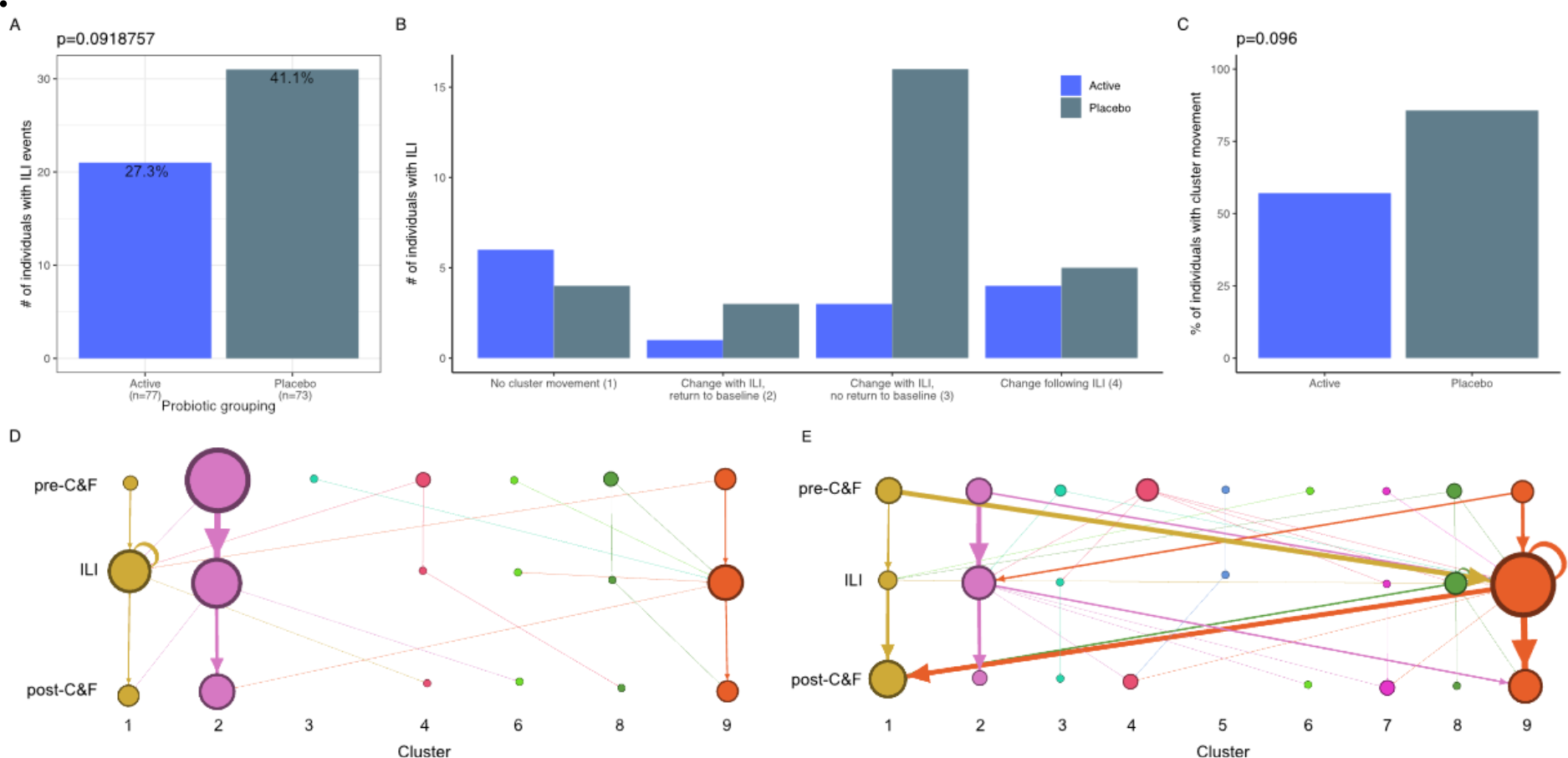
The effect of an oral *Lactobacillus rhamnosus GG* probiotic on ILI. A. There was no statistical difference in the number of individuals who experienced ILI between the probiotic (**active**) and placebo arms of the trial (p=0.0918757, chi-squared test). **B.** The number of participants in each of the 4 cluster movement categories separated by whether they were part of the probiotic/active or placebo arms of the trial. **C.** The frequency of cluster movement between individuals receiving probiotic vs. placebo treatments (p=0.096, chi-squared test). **D-E.** Network representation of cluster movement of individuals who experienced ILI on active (**D**) and placebo (**E**) treatment. Node size is proportional to the number of samples in a particular category/cluster group. The edges that connect node categories are sized proportional to the number of individuals who transitioned from one category to another during the course of their respiratory infection. Nodes and edges are coloured by the cluster that they belong to.

### The effect of influenza like illness (ILI) on the composition of the nasal microbiota

We compared the composition of the nasal microbiota when individuals had symptoms of ILI and when they did not. We found that the microbial community significantly differs between samples collected pre-/post-cold and influenza (C&F) season compared to during ILI (p=0.003, R^2^=0.05 (Bray Curtis); p=0.011, R^2^=0.04 (Aitchison) PERMANOVA; **Fig 2A**). These results are supported by another recent comparison of the effect of ILI on the frail, aged nasopharyngeal microbiota^36^. Alpha diversity, total bacterial load, and cluster membership were not altered between ILI and non-ILI samples (p=0.297, Levene’s test, **Fig 2B;** p=0.120, Levene’s test, **Fig 2C**; p=0.148, chi-squared test, **Sup Fig 5**). Further, there was no difference in the composition or cluster membership of the pre-C&F microbiota between those who went on to experience ILI and those who did not (p=0.378, R^2^=0.007 (Bray Curtis); p=0.095, R^2^=0.008 (Aitchison) PERMANOVA; **Sup Fig 6A;** p=0.121349, chi-squared test; **Sup Fig 6B**) indicating that we cannot predict who will get an infection based on the composition of the microbiota alone.

Given that the composition of the microbiome changes with ILI, we next investigated whether this change could be attributed to specific ASVs. Using ANCOMBC^29^, we identified 8 ASVs which were differentially abundant between the pre-C&F season, ILI, and post-C&F season; however, none of these has a mean relative abundance >0.1% (**Sup Fig 7**).

### A decrease in temporal stability of the nasal microbiota with ILI

Having determined that ILI affects the microbiome composition during illness, we next asked what effect ILI has following illness. Of those participants who reported symptoms of ILI during the study period, the composition of their microbiome before (pre-C&F) and after (post-C&F) ILI differed statistically from each other based on Aitchison (p=0.002, R^2^=0.015) but not Bray Curtis (p=0.267, R^2^=0.009) distance or cluster membership (p=0.528, chi-squared test). When the post-C&F composition of individuals who did and did not experience ILI were compared, the composition of the microbiome did not differ significantly (Aitchison, p=0.186, R^2^=0.009; Bray Curtis, p=0.095, R^2^=0.012; cluster membership, p=0.133, chi-squared test). Together, these results indicate that there is some effect of ILI on the microbiota following illness at the community level but that this effect is not consistent enough to distinguish between individuals who had and had not experienced these respiratory events.

When we analysed each individual – as opposed to focusing on community-wide metrics – we observe substantial changes to the microbiome during and following ILI. We tracked each individual’s microbiota across PCoA space and asked whether the rate of movement between clusters is affected by ILI. As shown previously, the dataset separates by cluster in a PCoA analysis; examining the chronological sampling of each individual, we see within individual movement across PCoA space (**Fig 3A**). By focusing on individuals who experienced ILI (n=51), we define 4 *movement categories*:

(1) individuals who stay in the same cluster before, during, and after ILI (**Fig 3B**); and individuals who move between clusters: (2) with ILI but later returning to the pre-ILI cluster (**Fig 3C**); (3) with ILI but not returning to the pre-ILI cluster (**Fig 3D**); and (4) following – but not during – ILI (**Fig 3E**). These categories do not correlate with collected metadata (**Sup Table 3**); however, the within individual mean Bray Curtis distance are increased in categories which resulted in a permanent change in cluster membership (3-4) when compared to individuals who did not change clusters (category 1; **Sup Fig 8)**, indicating the increased diversity between samples from individuals who experience significant cluster movement. 76.2% of individuals who experienced ILI moved between clusters, with 87.5% not returning to their original cluster by the end of the study period (categories 3-4; **Fig 3F**). Importantly, individuals who experienced ILI were statistically more likely to move between clusters when compared to those who did not have respiratory infection (76.2 vs. 48.7% of individuals; p=0.006, chi-squared test; **Fig 3G**). Movement between clusters was not predictable (e.g., there was no preference for a sample in a particular cluster to move to another at the next timepoint; **Sup Fig 9**). Together, these results indicate a significant legacy of change to the nasal microbiota associated with ILI events.

### The impact of probiotic use on ILI

The number of individuals experiencing ILI and the mean number of reported ILI events per individual did not differ statistically between the active and placebo arms of this study, as previously reported^17^ (p=0.092, chi-squared test, **Fig 4A**; p=0.589, t-test, data not shown). Neither the microbial composition nor cluster membership of individuals experiencing ILI differed statistically between probiotic and placebo treatments (p=0.438, R^2^=0.018 (Bray Curtis); p=0.529, R^2^=0.017, (Aitchison) distances; p=0.220, chi-squared test). Similarly, the post-C&F microbiota did not differ statistically with probiotic use (p=0.458, R^2^=0.008 (Bray Curtis); p=0.809, R^2^=0.007 (Aitchison) distances; p=0.178, chi-squared test). Cluster membership nor microbiome composition were affected by the probiotic itself (**Sup Fig 10**). Collectively, this indicates that probiotic use does not affect the microbial composition of the nose during or following ILI in community-wide analyses.

In contrast, individuals who experienced ILI whilst receiving probiotic were less likely to move between clusters compared to those experiencing ILI on placebo treatment (**Fig 4B**); 24 (85.7%) of individuals on placebo treatment moved between clusters in comparison to 8 (57.1%) individuals on probiotics (**Fig 4C**). This observation is not statistically significant (p=0.096, chi-squared test); further investigation of a larger cohort is needed to determine whether probiotic use could significantly affect the stability of the microbiome. In particular, there was more movement between clusters 1 and 9 at the onset/resolution of ILI in individuals treated with placebo versus probiotic (**Fig 4D-E).** Together, these results indicate that the administration of probiotics did not have an observable effect on the overall nasal microbiota and that further studies are needed to assess whether probiotics can mitigate the long-term impact of ILI on the individual.

## Discussion

Here we show that the diverse microbiota of frail, older residents of long-term care homes could be grouped into 9 distinct clusters based on ASV presence and abundance. We find that the nasal microbiota of frail older adults exhibits an individualized response to ILI, often resulting in a lack of stability which is possibly mitigated, at least in part, by probiotic use.

The observed diversity of this community is perhaps not surprising given that inter-individual variability is also a feature of the aging immune system, where a lifetime of environmental exposures and immune experiences shapes the immune response and age-associated inflammation^37^. Interestingly, age did not correlate with microbiota composition, but frailty did, results which are in line with frailty being a better indicator of infection risk than age^4, 5^.

Although this study is unique in its focus on a more frail, LTC dwelling cohort, previous studies have similarly identified changes to the respiratory tract microbiota in individuals who experience respiratory infection^38, 39^. In particular, a recent study of older (mean age of 70) community dwelling adults found similar distinctions between individuals experiencing ILI versus healthy controls^36^. Interestingly, this study found a difference in the stability of the microbiota post-ILI in individuals with higher abundances of core microbiota species (including *Corynebacterium, Dolosigranulum,* and *Staphylococcus*) ^36^; in contrast, we see no evidence of a difference in stability between individuals in clusters associated with or without a dominant taxa, perhaps suggesting that any protective effect of dominant taxa from ILI-induced changes to the microbiota in healthy older adults is weaker in this frail population.

We identified a microbiome that lacked stability and changed longitudinally in 76% of individuals with ILI. The microbiota did not change in a predictable way or converge on a particular ASV or cluster, but instead was highly individualized. These results indicate that the introduction of a pathogen – be it viral or bacterial – often leads to profound changes in the frail, aged nasal microbiota. The results of the pilot study of a probiotic targeted for the gastrointestinal tract suggest that it may be possible to mitigate these changes, at least in part. Only 21 (of 77) individuals on probiotic treatment experienced ILI, and of those only 8 moved between microbiome clusters; this is in contrast to 30 of 73 individuals on placebo experiencing ILI with 24 of those 30 moving between clusters. This pilot trial is too small to be able to statistically conclude that probiotic use is beneficial to the stability of the nasal microbiota during and following ILI, and further investigation – perhaps with a nasal probiotic – are encouraged.

Our analyses identify 9 distinct clusters of nasal microbial communities across this dataset. All but one of these clusters are associated with a dominant taxa (present at >35% relative abundance in >70% of samples), similar to that of the, community dwelling older adult microbiota^36^. Our use of hierarchical clustering outlined the 4 ASVs of *Corynebacterium* and 5 ASVs of *Moraxella* prevalent in the dataset. The ASVs of each species rarely co-occur within an individual (with the exception of cluster 1) suggesting possible intra-species competition within this niche. This may have downstream implications on the ecology of these communities, especially when we consider that certain *Corynebacterium* and *Moraxella* ASVs were differentially abundant across various collected metadata (**Sup Fig 4**). Cluster 2 was unique in that it wasn’t dominated by a particular taxa and that it was more diverse than the other clusters as measured by the Shannon diversity index. Interestingly, this increase in diversity correlated with a decrease in bacterial load (as measured by qPCR concentration), perhaps indicating the loss of a once-present prevalent taxon leaving only the less abundant – but highly diverse – taxa in its wake.

Of the tested metadata variables, we identified a correlation of microbial composition with LTC home site, sex, time of collection, frailty, and cardiovascular disease. Variability of the microbiota with LTC home is already well-established in the gut^16^ and underlies known variations in management practices, air quality, diet, location etc. between LTC home sites. Similarly, changes to the nasal microbiota with the seasons has been previously documented in children^40^, and year-to-year differences may represent the effect of circulating viruses (and variants thereof) on the nasal microbiota.

Preventing respiratory infection – and/or the long-term consequences of – in frail older adults will have an outsized impact on their care, quality of life and use of health care resources. Frailty, disability, and loss of independence is exacerbated by having a respiratory infection^41^ and hospitalization rates – especially for strokes and cardiorespiratory events – increase months to years after infection^42^. Some, but not all vaccines, are less effective in frail individuals^43^ so understanding the features of the frail microbiome and exploring new preventative measures are essential to reducing the burden of respiratory infections.

## Declarations

### Ethics approval and consent to participate

Informed consent was received by the participants or their substitute decision makers. All protocols were approved by the Hamilton Integrated Research Ethics Board.

### Consent for publication

Not applicable.

### Availability of data and materials

All sequencing data is publicly available in NCBI’s SRA PRJN858212. All code used to generate analyses presented in all Tables/the text and all visuals presented in Figures is available in an R markdown file (**Sup File 1**).

### Author Contributions

FJW is the primary author of this manuscript; FJW processed some and analyzed all data. MF conducted DNA extractions of samples. LR conducted all PCR reactions and post-sequencing processing of the data. LPS, and JT conducted the qPCR measurements. FJW, DMEB, MGS, JJ, and ML conceptualized the experimental outline. FJW wrote the manuscript. All authors edited and approved the manuscript.

## Data Availability

All data produced in the present work are contained in the manuscript or online at locations detailed in the manuscript and/or its supplemental material.

## Acknowledgements

We wish to acknowledge the individuals who agreed to take part in this study without which these results would not be possible.

## Supplemental Materials

**Supplemental Figure 1:**
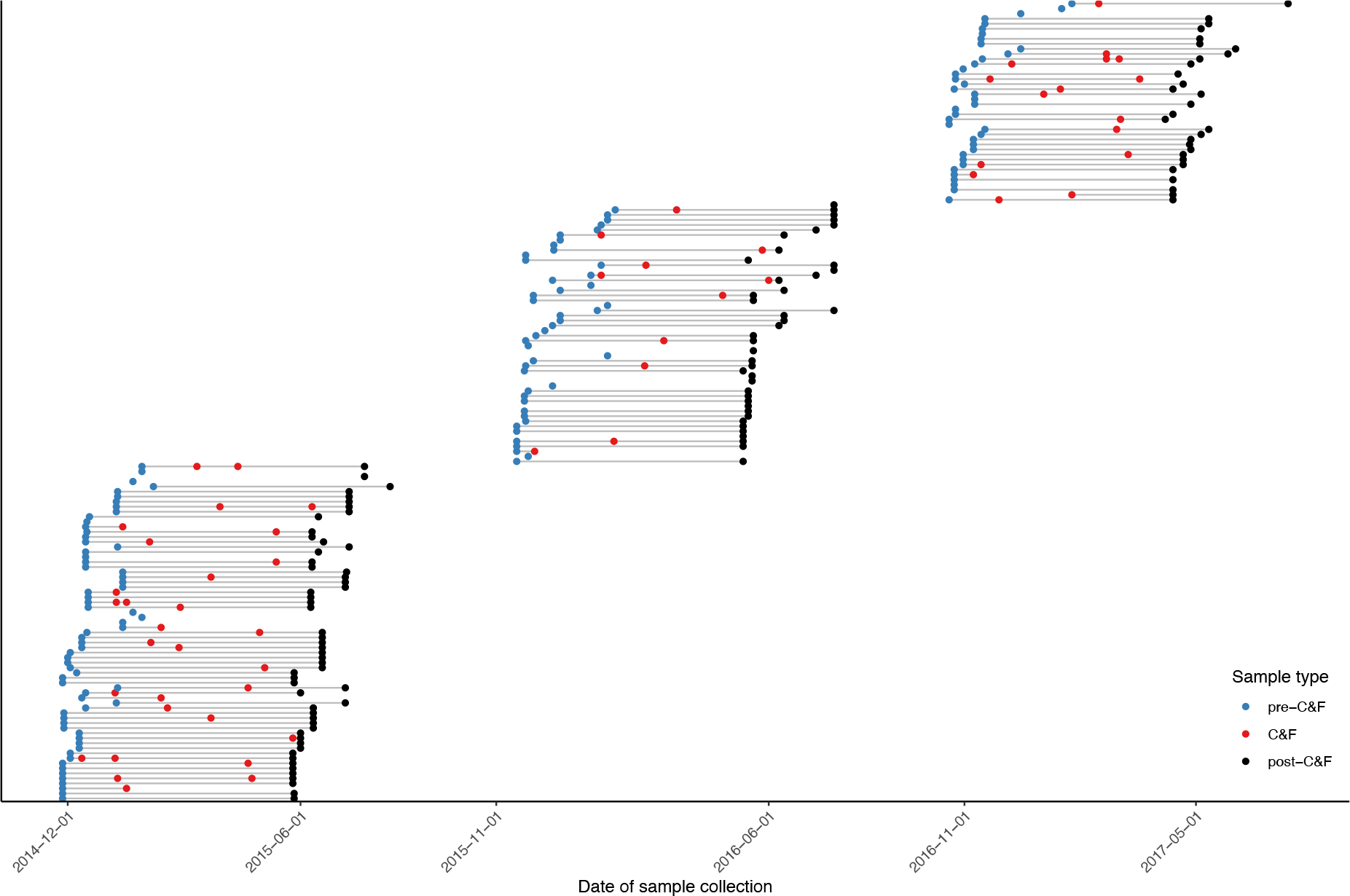
A time series schematic of mid-turbinate sample collection. A time series of sample collection in this multi-year, longitudinal dataset (n=334). Labelled dates on the x-axis represent approximate start and end dates of collection for each cohort of samples. pre-C&F = pre-cold and influenza season; post-C&F = post-cold and influenza season; ILI = influenza like illness.

**Supplemental Figure 2:**
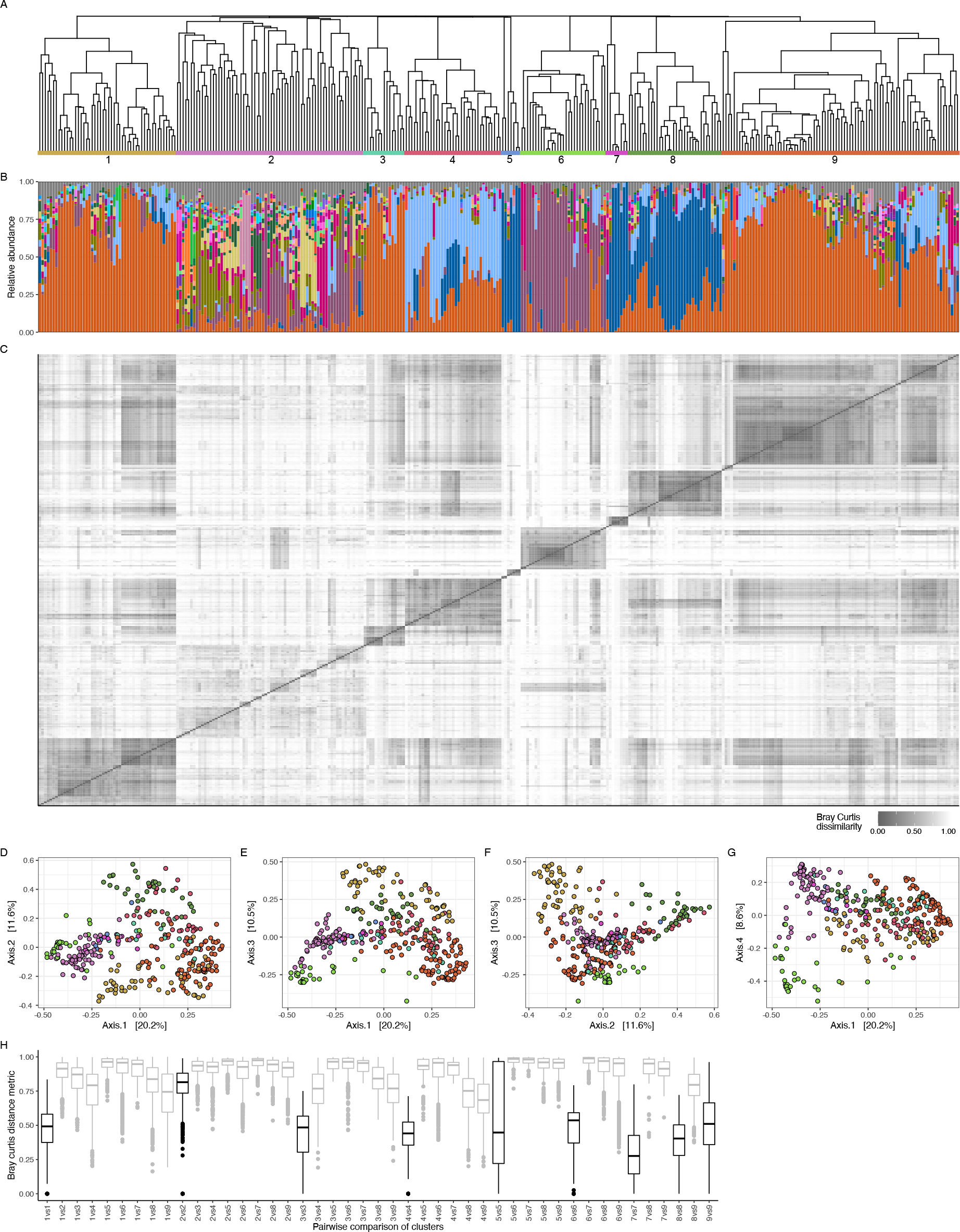
**Evidence that 9 clusters is optimal. A-B**. The dendrogram and taxonomic summaries as shown in Fig 1 redisplayed here for comparison. A detailed legend matching colours to genus-level taxonomic assignments is provided in **Sup** Fig 3. **C.** A heatmap of Bray Curtis distances between samples ordered as in **panels A-B**. **D-G**. PCoA analyses of various axes show separation of samples from each cluster across PCoA space. **H**. Median Bray Curtis distances within (**black**) and between (**grey**) each cluster show more similarity within clusters (with the exception of cluster 2) than between clusters.

**Supplemental Figure 3:**
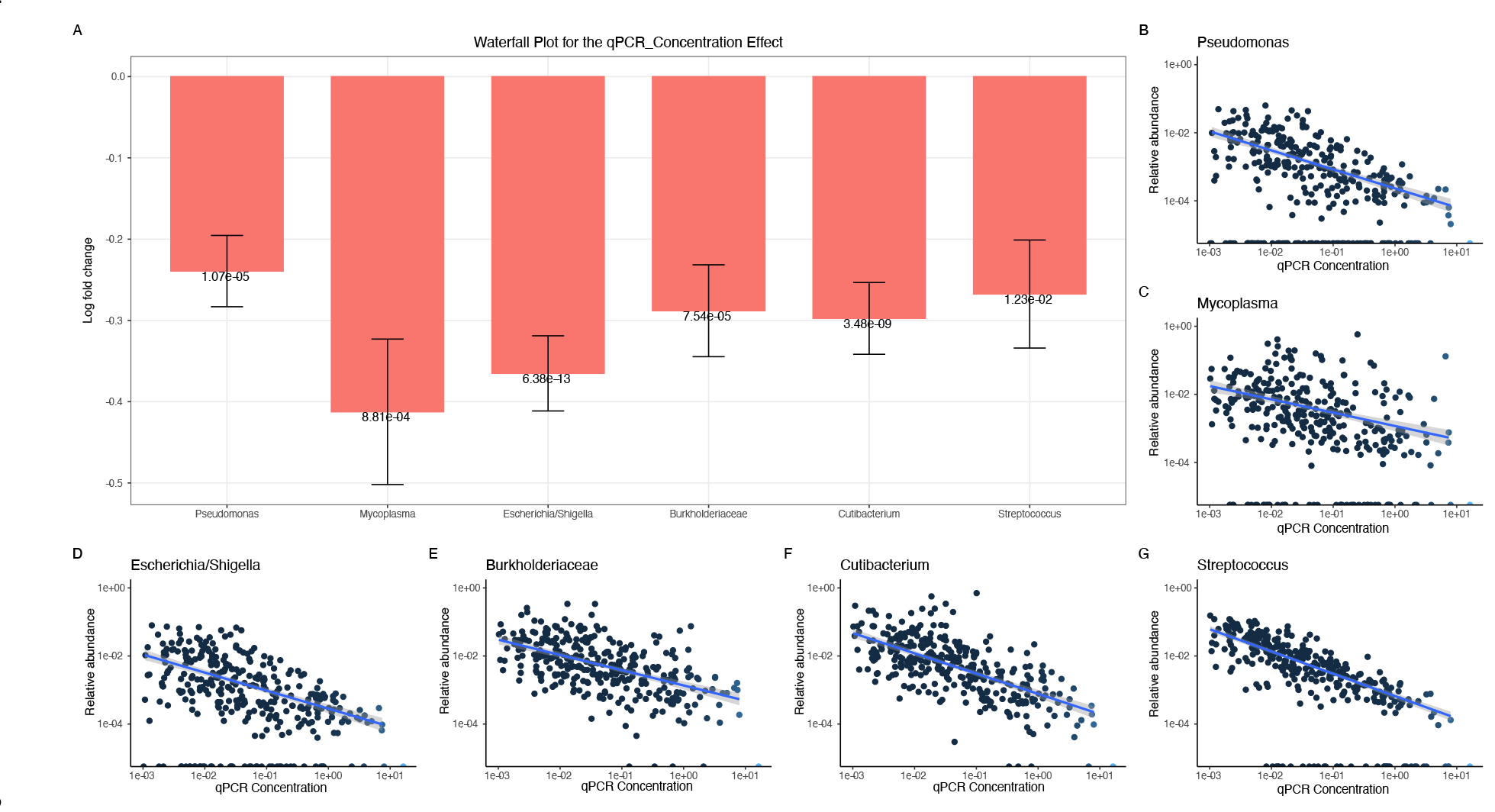
ASVs which are differentially abundant with qPCR concentration. A. ANCOMBC was used to determine ASVs which were differentially abundant with qPCR concentration. Here, the log fold change of each differentially abundant ASV is displayed as well as the adjusted p-value. The genus (or family if the genus was undefined) taxonomic id is used to identify each ASV. **B-G.** The log-transformed qPCR concentration and relative abundance of each differentially abundant ASV is shown. A regression line with confidence intervals is shown in **blue.**

**Supplemental Figure 4:**
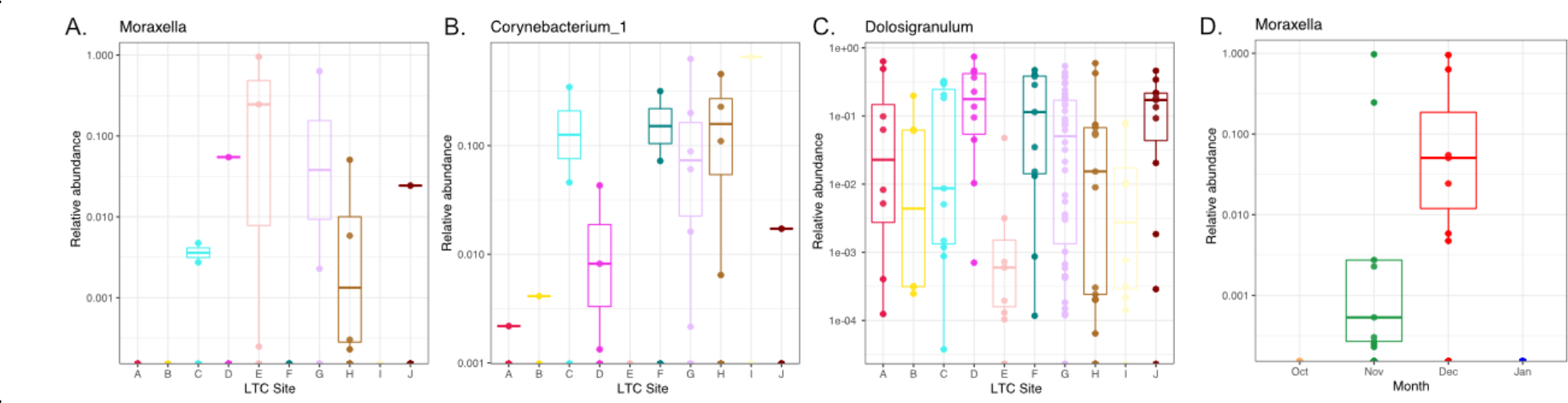
ASVs which significantly correlate with particular metadata variables. A-C. When examining the LTC site, among the 9 differentially abundant ASVs with a mean relative abundance >0.1% include 3 taxa which are dominant across the dataset. **D.** Of the 5 differentially abundant ASVs across the month of collection, 2 also correlated with LTC site, including a *Moraxella* ASV.

**Supplemental Figure 5:**
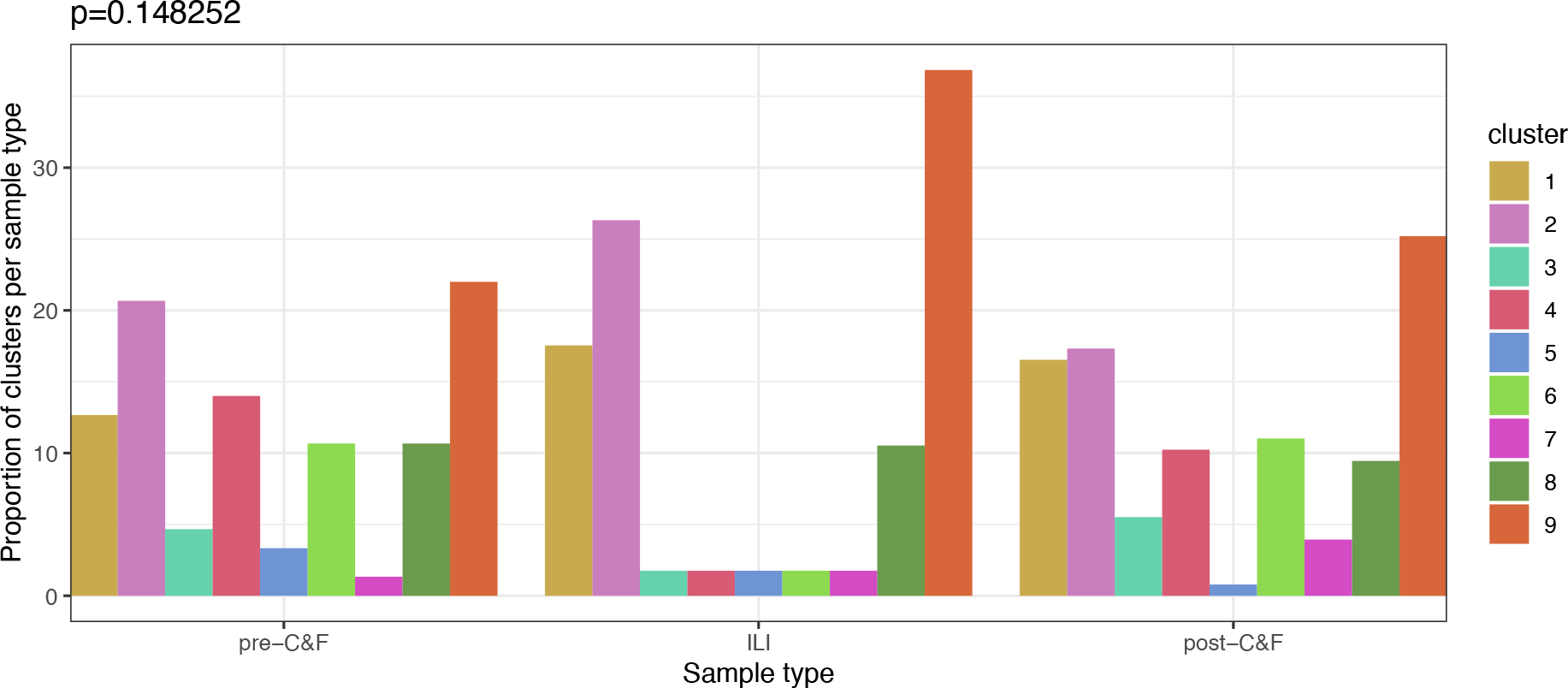
**The proportion of pre-C&F, ILI, and post-C&F samples in each cluster type**. Samples collected at different points in the study period were not preferentially found in any particular cluster (p=0.148252, chi-squared test).

**Supplemental Figure 6:**
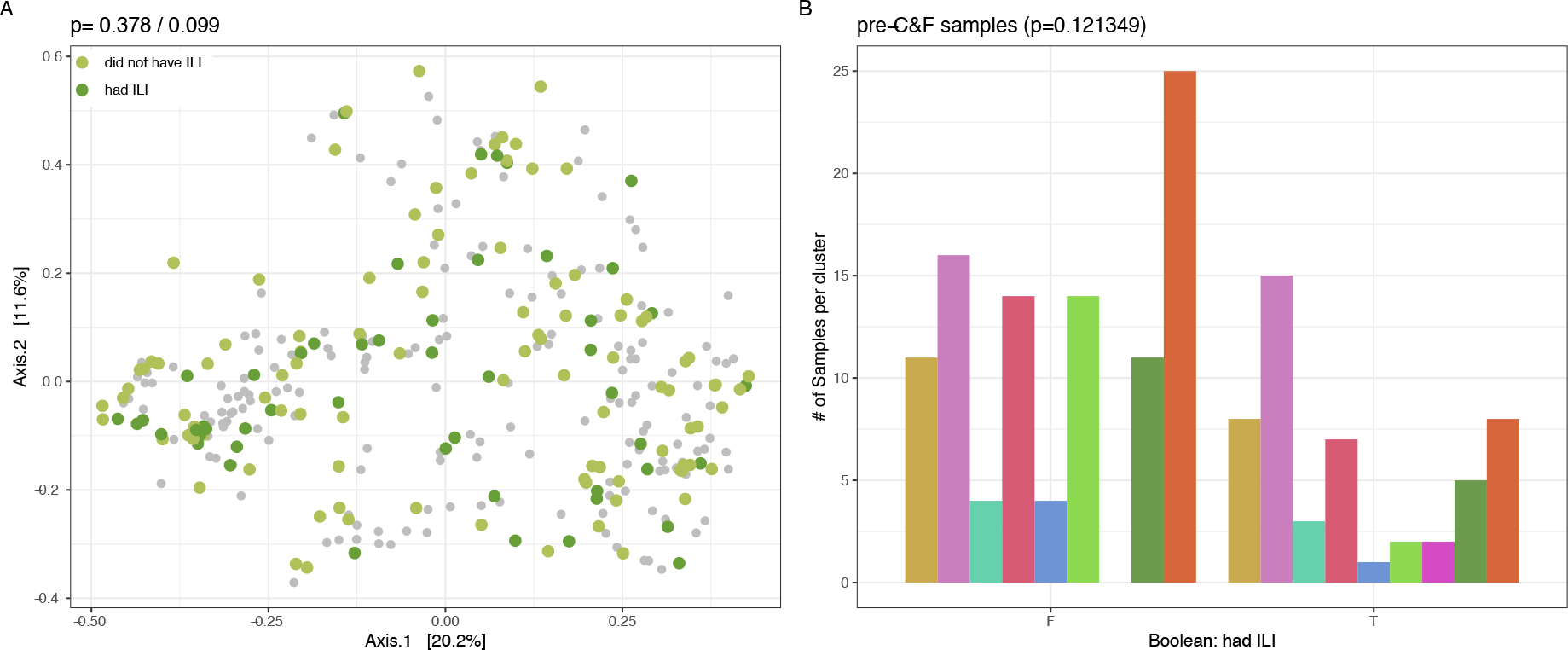
Respiratory events are not predictable from *a priori* collected samples. A. There are no observable differences between pre-C&F samples collected before ILI events did or did not occur (p=0.378/0.099, permanova with Bray Curtis and Aitchison distances, respectively). **B.** Clustering of pre-C&F samples did not statistically differ between individuals who did and did not subsequently experience ILI (p=0.121349, chi-squared test).

**Supplemental Figure 7:**
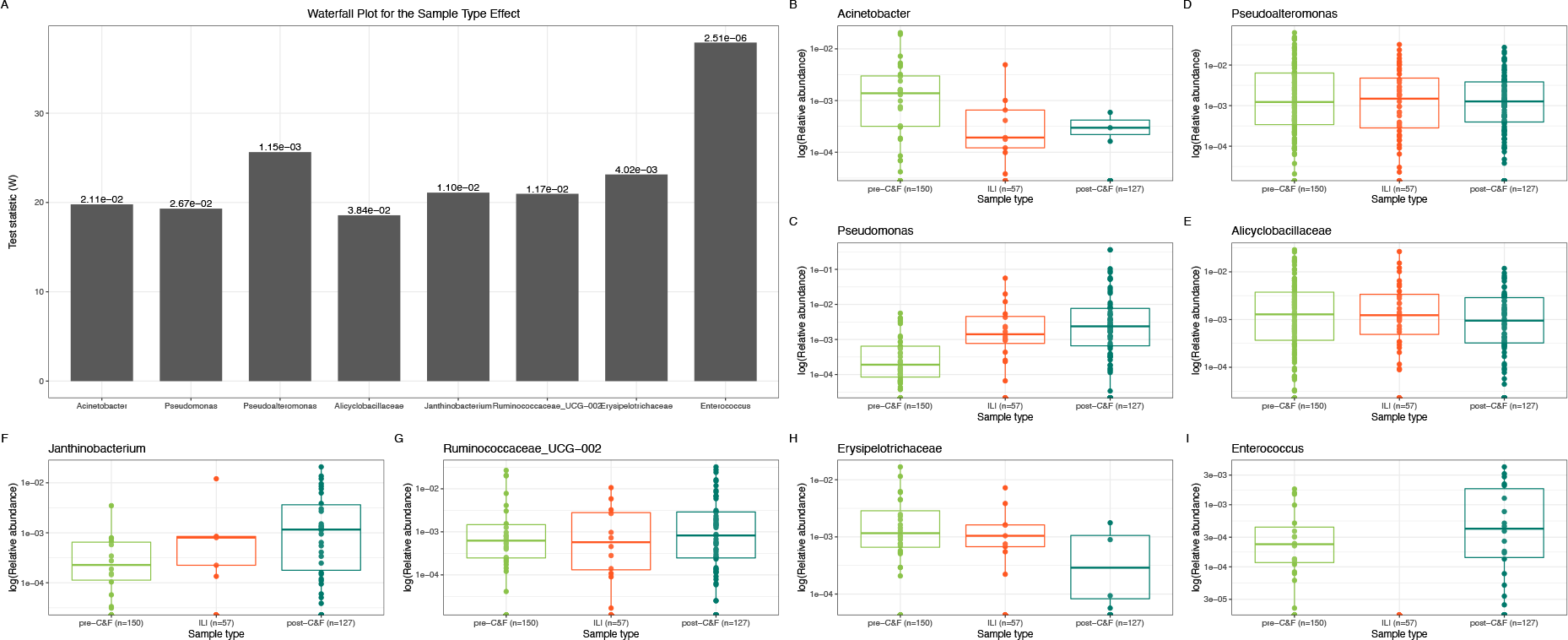
ASVs which are differentially abundant across pre-C&F, ILI, and post-C&F sample types. A. ANCOMBC was used to determine the ASVs which were differentially abundant across the three sample types. Here, the test statistic (W) of each differentially abundant ASV is displayed along with the adjusted p-value. The genus (or family if the genus was undefined) taxonomic ID is used to identify each ASV. **B-I.** The log-transformed relative abundance of each differentially abundant ASV across the three sample types.

**Supplemental Figure 8:**
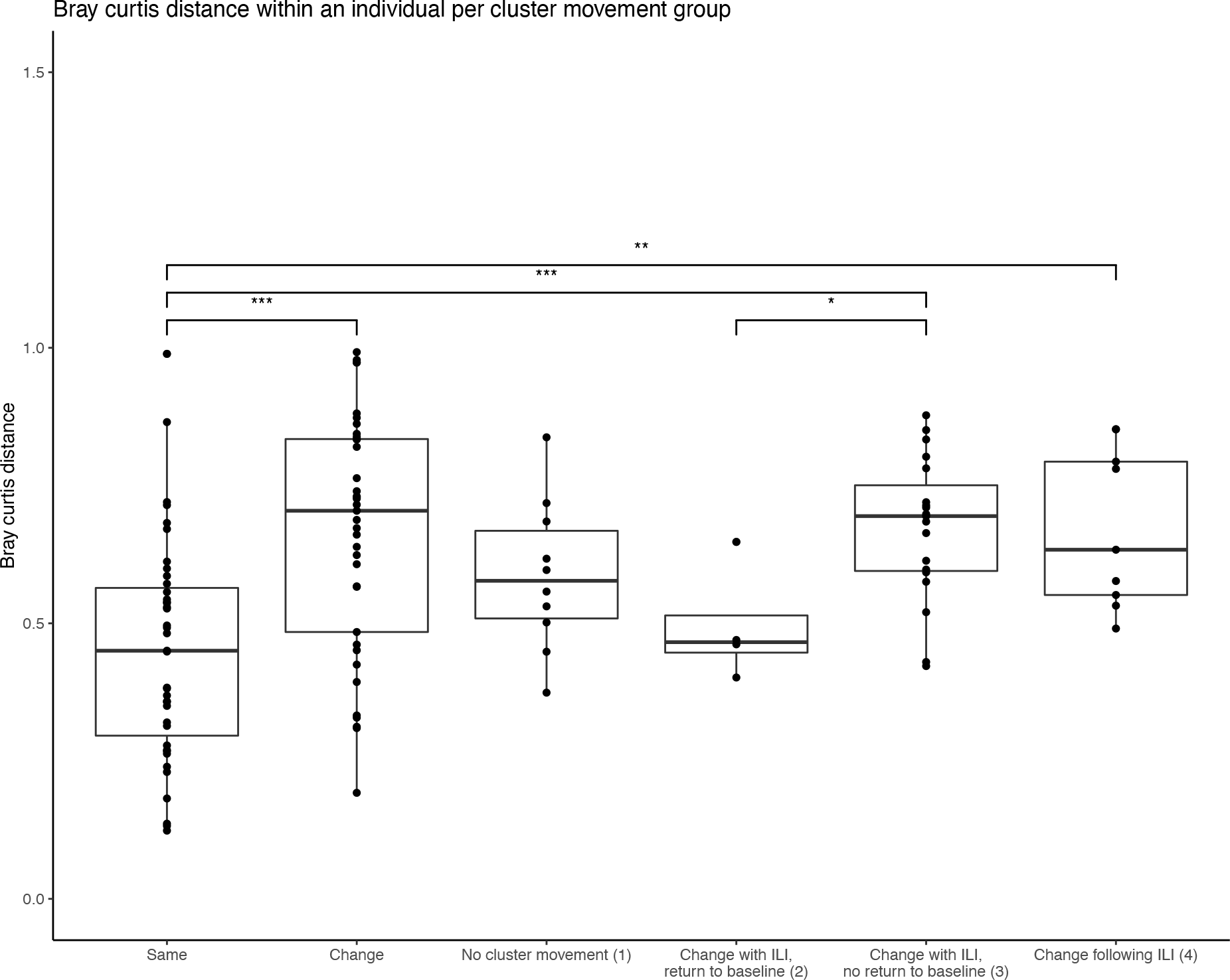
The intra-individual Bray Curtis distance is increased when significant cluster movement is observed. **In individuals who did not have** events, those whose microbiome moved between clusters (**“change”**) have a statistically significant increased Bray Curtis distance when compared to those who did not move between clusters (**“same”**). Similarly, individuals who experienced influenza-like illness (ILI) had an increased intra-individual Bray Curtis distance when the illness resulted in a permanent change in cluster membership (categories 3-4) when compared to individuals who did not change clusters (category 1).

**Supplemental Figure 9:**
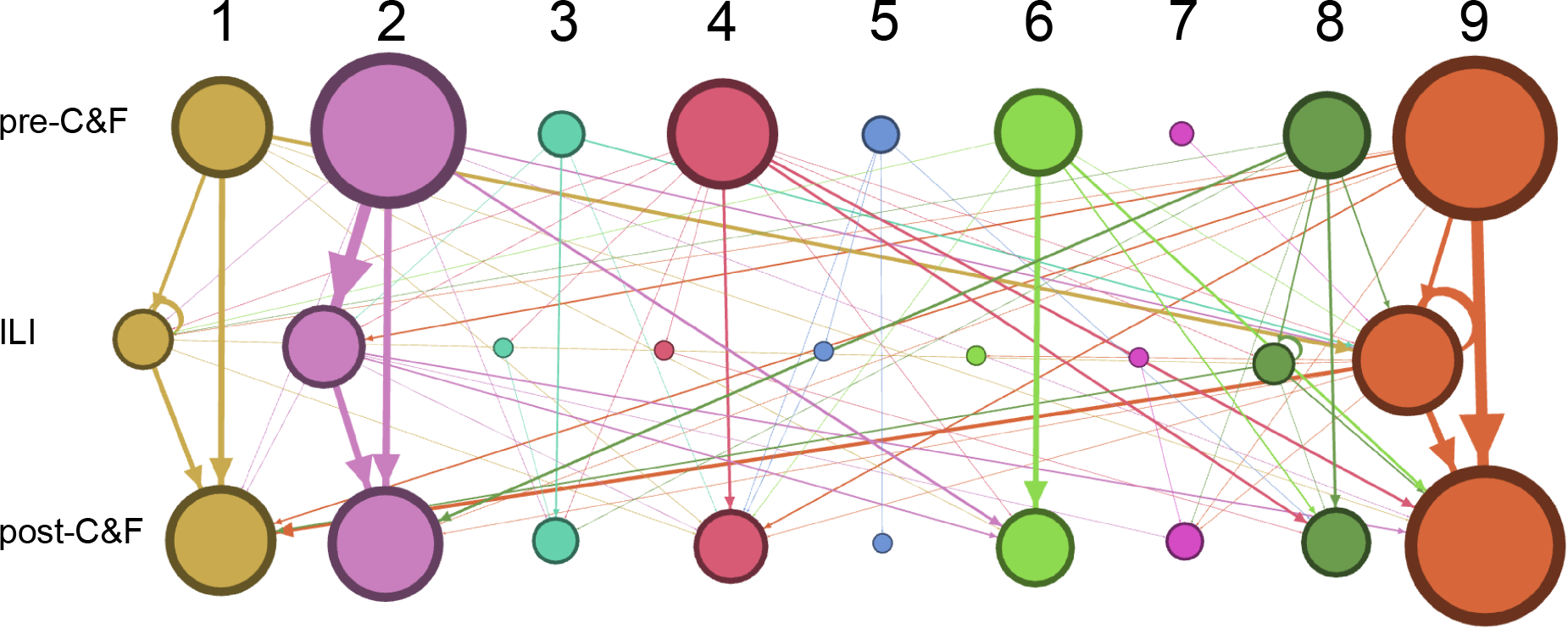
There is no discernible pattern in cluster movement between samples. Nodes and edges are weighted based on the number of samples in each category. Rows and columns of nodes are labelled with the sample type (pre- C&F, ILI, post-C&F) and cluster number (1-9).

**Supplemental Figure 10:**
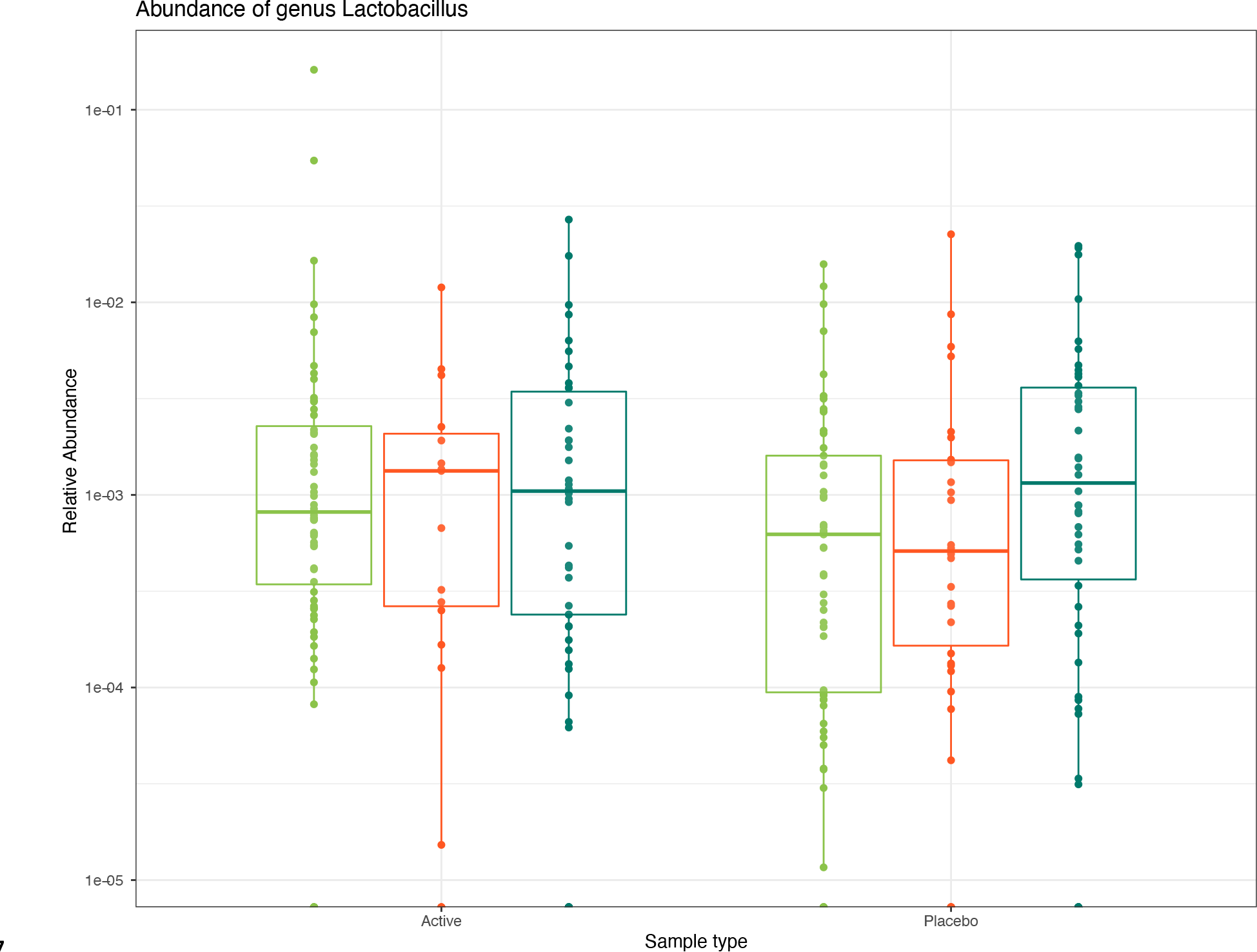
Relative abundance of *Lactobacillus* across samples. There were no differentially abundant ASVs between individuals on active and placebo treatments, including any ASV with the taxonomic assignment of *Lactobacillus*.

**Supplemental Table 1:**
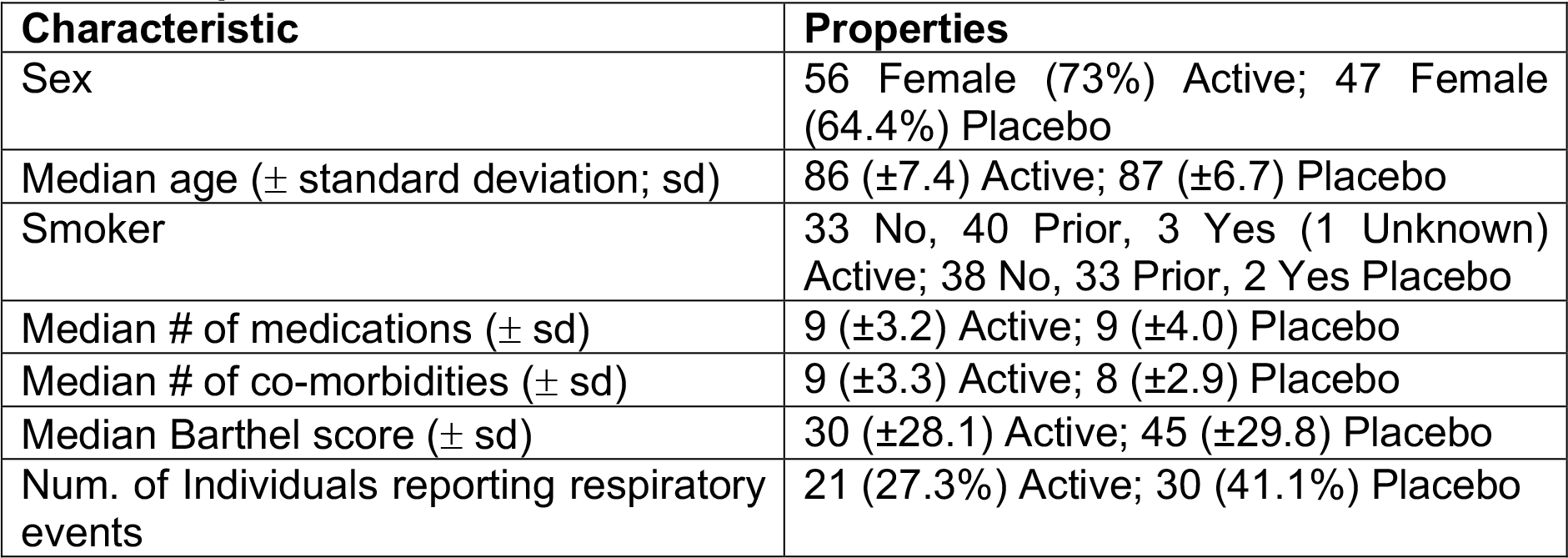
Participant characteristics across the dataset split by active and placebo arms of the trial.

**Supplemental Table 2:** Correlating metadata variables.

| **Variable 1** | **Variable 2** | **Statistical test, p-value** |
| --- | --- | --- |
| Age at enrolment | Sex | aov, p=0.040 |
| Age at enrolment | Smoker | aov, p=0.0002 |
| Age at enrolment | Influenza vaccination (current season) | aov, p=0.044 |
| Age at enrolment | Anemia | aov, p=0.006 |
| Age at enrolment | Seizures | aov, p=0.020 |
| Site at enrolment | Month | chisq, p=2.519e-05 |
| LTC Site | Season | chisq, p=0.009 |
| LTC Site | Year | chisq, p=2.427e-13 |
| LTC Site | Influenza vaccination (previous season) | chisq, p=0.026 |
| LTC Site | Pneumococcal vaccine (ever) | chisq, p=0.001 |
| LTC Site | Shared room | chisq, p=9.711e-10 |
| LTC Site | Seizures | chisq, p=0.041 |
| Month | Season | chisq, p < 2.2e-16 |
| Month | Year | chisq, p=7.831e-16 |
| Month | Smoker | chisq, p=0.0004 |
| Month | Pneumococcal vaccine (ever) | chisq, p=0.024 |
| Month | IL1B | aov, p=0.005 |
| Season | Year | chisq, p=0.0003 |
| Season | Pneumococcal vaccine (ever) | chisq, p=0.019 |
| Season | Dementia | chisq, p=0.018 |
| Season | IL1B | aov, p=0.027 |
| Had respiratory event | IL1B | aov, p=0.042 |
| Smoker | COPD | chisq, p=0.012 |
| Smoker | IL1B | aov, p=0.050 |
| Medications (number of) | COPD | aov, p=0.015 |
| Medications (number of) | CHF | aov, p=0.0003 |
| Medications (number of) | Dementia | aov, p=0.017 |
| Medications (number of) | DM | aov, p=0.022 |
| Medications (number of) | Comorbidities (number of) | aov, p=8.03e-07 |
| Influenza vaccination (current season) | Influenza vaccination (previous season) | chisq, p=0.001 |
| Influenza vaccination (current season) | Influenza vaccination (ever) | chisq, p=3.596e-08 |
| Influenza vaccination (current season) | Pneumococcal vaccine (ever) | chisq, p=7.083e-05 |
| Influenza vaccination (current season) | IL1B | aov, p=0.032 |
| Influenza vaccination (previous season) | Influenza vaccination (ever) | chisq, p=7.785e-10 |
| Influenza vaccination (previous season) | Pneumococcal vaccine (ever) | chisq, p=0.001 |
| Influenza vaccination (previous season) | Comorbidities (number of) | aov, p=0.014 |
| Influenza vaccination (ever) | Pneumococcal vaccine (ever) | chisq, p=0.003 |
| Influenza vaccination (ever) | Barthel total | aov, p=0.037 |
| Influenza vaccination (ever) | Comorbidities (number of) | aov, p=0.044 |
| Influenza vaccination (ever) | IL1B | aov, p=0.015 |
| Pneumococcal vaccine (ever) | Cancer | chisq, p= 0.029 |
| COPD | CVD | chisq, p=0.002 |
| CHF | Comorbidities (number of) | aov, p=0.002 |
| CVD | Comorbidities (number of) | aov, p=2.56e-06 |
| Anemia | Comorbidities (number of) | aov,p= 0.008 |
| Dementia | IL6 | aov, p=0.027 |
| Stroke | Comorbidities (number of) | aov, p=0.017 |
| Diabetes mellitus | Comorbidities (number of) | aov, p=0.045 |

**Supplemental Table 3:** p-values of correlation tests (per individual) between collected metadata variables and cluster movement categories.

| Characteristic | p-value | Statistical test |
| --- | --- | --- |
| Age at enrolment | 0.312 | aov |
| Sex | 0.123 | chisq |
| LTC home site | 0.722 | chisq |
| Allocation Group<br>Probiotics | 0.091 | chisq |
| Smoker | 0.192 | chisq |
| Num Medications | 0.326 | aov |
| Influenza vacc this season | 0.743 | aov |
| Influenza vacc last season | 0.419 | chisq |
| Influenza vaccine ever | 0.447 | chisq |
| Has pt received pneumonia vaccine | 0.290 | chisq |
| Is pt in shared room | 0.985 | chisq |
| Barthel total | 0.668 | aov |
| COPD | 0.190 | chisq |
| CHF | 0.148 | chisq |
| CVD | 0.424 | chisq |
| Anemia | 0.206 | chisq |
| Dementia | 0.312 | chisq |
| CVA Stroke | 0.123 | chisq |
| DM | 0.403 | chisq |
| Hypothyroid | 0.176 | chisq |
| Num Comorbidities | 0.725 | aov |
| Seizures | 0.309 | chisq |
| Cancer | 0.468 | chisq |
| IL1B | 0.438 | aov |
| IL6 | 0.569 | aov |
| TNFA | 0.675 | aov |

**Correlation was determined either with a chi squared or aov test, depending on the data type of the variable. COPD: chronic obstructive pulmonary disease, CHF: congestive heart failure, CVD: cardiovascular disease.**

## References

1. Childs, A., et al., The burden of respiratory infections among older adults in long-term care: a systematic review. BMC Geriatrics, 2019. 19(1): p. 210.

2. Bosco, E., et al., Long-term Care Facility Variation in the Incidence of Pneumonia and Influenza. Open forum infectious diseases, 2019. 6(6): p. ofz230-ofz230.

3. Brown, K.A., et al., Association Between Nursing Home Crowding and COVID-19 Infection and Mortality in Ontario, Canada. JAMA Internal Medicine, 2021. 181(2): p. 229–236.

4. Iwai-Saito, K., et al., Frailty is associated with susceptibility and severity of pneumonia in older adults (A JAGES multilevel cross-sectional study). Scientific Reports, 2021. 11(1): p. 7966.

5. Grabovac, I., et al., Frailty Status Predicts All-Cause and Cause-Specific Mortality in Community Dwelling Older Adults. Journal of the American Medical Directors Association, 2019. 20(10): p. 1230–1235.e2.

6. Bleve, A., et al., Immunosenescence, Inflammaging, and Frailty: Role of Myeloid Cells in Age-Related Diseases. Clin Rev Allergy Immunol, 2022: p. 1–22.

7. Butler, J.C. and A. Schuchat, Epidemiology of Pneumococcal Infections in the Elderly. Drugs & Aging, 1999. 15(1): p. 11–19.

8. Flamaing, J., et al., Pneumococcal colonization in older persons in a nonoutbreak setting. J Am Geriatr Soc, 2010. 58(2): p. 396–8.

9. Mitsi, E., et al., Nasal Pneumococcal Density Is Associated with Microaspiration and Heightened Human Alveolar Macrophage Responsiveness to Bacterial Pathogens. Am J Respir Crit Care Med, 2020. 201(3): p. 335–347.

10. de Steenhuijsen Piters, W.A.A., et al., Interaction between the nasal microbiota and S. pneumoniae in the context of live-attenuated influenza vaccine. Nature Communications, 2019. 10(1): p. 2981.

11. Xu, L., J. Earl, and M.E. Pichichero, Nasopharyngeal microbiome composition associated with Streptococcus pneumoniae colonization suggests a protective role of Corynebacterium in young children. PLoS One, 2021. 16(9): p. e0257207.

12. Henares, D., et al., Differential nasopharyngeal microbiota composition in children according to respiratory health status. Microb Genom, 2021. 7(10).

13. Man, W.H., et al., Bacterial and viral respiratory tract microbiota and host characteristics in children with lower respiratory tract infections: a matched case-control study. The Lancet Respiratory Medicine, 2019. 7(5): p. 417–426.

14. de Steenhuijsen Piters, W.A.A., et al., Dysbiosis of upper respiratory tract microbiota in elderly pneumonia patients. The ISME Journal, 2016. 10(1): p. 97–108.

15. DeJong, E.N., M.G. Surette, and D.M.E. Bowdish, The Gut Microbiota and Unhealthy Aging: Disentangling Cause from Consequence. Cell Host Microbe, 2020. 28(2): p. 180–189.

16. Claesson, M.J., et al., Gut microbiota composition correlates with diet and health in the elderly. Nature, 2012. 488(7410): p. 178–84.

17. Wang, B., et al., Probiotics to Prevent Respiratory Infections in Nursing Homes: A Pilot Randomized Controlled Trial. J Am Geriatr Soc, 2018. 66(7): p. 1346–1352.

18. Whelan, F.J., et al., The loss of topography in the microbial communities of the upper respiratory tract in the elderly. Ann Am Thorac Soc, 2014. 11(4): p. 513–21.

19. Bartram, A.K., et al., Generation of multimillion-sequence 16S rRNA gene libraries from complex microbial communities by assembling paired-end illumina reads. Applied and Environmental Microbiology, 2011. 77(11): p. 3846–52.

20. Martin, M., Cutadapt removes adapter sequences from high-throughput sequencing reads. EMBnet.journal. 17.

21. Callahan, B.J., et al., DADA2: High-resolution sample inference from Illumina amplicon data. Nat Methods, 2016. 13(7): p. 581–3.

22. Quast, C., et al., The SILVA ribosomal RNA gene database project: improved data processing and web-based tools. Nucleic Acids Res, 2013. 41(Database issue): p. D590- 6.

23. Lamarche, D., et al., Microbial dysbiosis and mortality during mechanical ventilation: a prospective observational study. Respiratory Research, 2018. 19(245).

24. Bacchetti De Gregoris, T., et al., Improvement of phylum- and class-specific primers for real-time PCR quantification of bacterial taxa. J Microbiol Methods, 2011. 86(3): p. 351–6.

25. Team, R.C., R: A language and environment for statistical computing. R Foundation for Statistical Computing, Vienna, Austria. 2020.

26. McMurdie, P.J. and S. Holmes, phyloseq: an R package for reproducible interactive analysis and graphics of microbiome census data. PLoS One, 2013. 8(4): p. e61217.

27. Oksanen, J., et al., vegan: Community Ecology Package. R package version 2.5-7. 2020.

28. Lahti, L. and S. Shetty, Tools for microbiome analysis in R. 2017.

29. Lin, H. and S.D. Peddada, Analysis of compositions of microbiomes with bias correction. Nat Commun, 2020. 11(1): p. 3514.

30. Bastian, M. and S. Heymann, Gephi An open source software for exploring and manipulating networks. International AAAI Conference on Weblogs and Social Media, 2009.

31. Wickham, H., ggplot2: Elegant Graphics for Data Analysis., S.-V.N. York, Editor. 2016.

32. Kassambara, A. and F. Mundt, Extract and Visualize the Results of Multivariate Data Analyses. 2020.

33. Maechler, M., et al., cluster: Cluster Analysis Basics and Extensions. R package version 2.*1*.*2*. 2021.

34. Yu, G., et al., ggtree: an R package for visualization and annotation of phylogenetic trees with their covariates and other associated data. Methods in Ecology and Evolution, 2017. 8(1): p. 28–36.

35. Mahoney, F.I. and D.W. Barthel, Functional Evaluation: The Barthel Index. Md State Med J, 1965. 14: p. 61–5.

36. Shetty, S.A., et al., Associations and recovery dynamics of the nasopharyngeal microbiota during influenza-like illness in the aging population. Sci Rep, 2022. 12(1): p. 1915.

37. Thevaranjan, N., et al., Age-Associated Microbial Dysbiosis Promotes Intestinal Permeability, Systemic Inflammation, and Macrophage Dysfunction. Cell Host Microbe, 2017. 21(4): p. 455–466 e4.

38. Kaul, D., et al., Microbiome disturbance and resilience dynamics of the upper respiratory tract during influenza A virus infection. Nat Commun, 2020. 11(1): p. 2537.

39. Ding, T., et al., Microbial Composition of the Human Nasopharynx Varies According to Influenza Virus Type and Vaccination Status. 2019.

40. Bogaert, D., et al., Variability and diversity of nasopharyngeal microbiota in children: a metagenomic analysis. PLoS One, 2011. 6(2): p. e17035.

41. Lees, C., et al., Frailty Hinders Recovery From Influenza and Acute Respiratory Illness in Older Adults. The Journal of Infectious Diseases, 2020. 222(3): p. 428–437.

42. Bosco, E., et al., Estimated Cardiorespiratory Hospitalizations Attributable to Influenza and Respiratory Syncytial Virus Among Long-term Care Facility Residents. JAMA Network Open, 2021. 4(6): p. e2111806–e2111806.

43. Yao, X., et al., Frailty is associated with impairment of vaccine-induced antibody response and increase in post-vaccination influenza infection in community-dwelling older adults. Vaccine, 2011. 29(31): p. 5015–5021.

